# Mapping Inter-Metabolite Relationships in the Human Brain: A Multi-Site, Multi-Modal MRS Study

**DOI:** 10.1101/2025.11.17.25340250

**Authors:** Ramona Leenings, Irene Neuner, Ravichandran Rajkumar, N. Jon Shah, Ezequiel Farrher, Daniel Güllmar, Ralf Mekle, Ivana Galinovic, Gilbert Hangel, Wolfgang Bogner, Philipp Lazen, Jonathan Mathias Fasshauer, Luise Victoria Claaß, Sarah Mackert, Carmen Uckermark, Marius Gruber, Jochen Bauer, Jonathan Repple, Nils C. Gassen, Sharmili Edwin Thanarajah, Alexander Gussew, Nils Opel

## Abstract

Technological advances in magnetic resonance spectroscopy (MRS), particularly at ultra-high field strengths (e.g., 7T), now allow for the reliable quantification of an increasing number of brain metabolites across multiple brain regions. However, most studies remain constrained to analyses of isolated metabolite levels, overlooking the complex interdependencies that govern neurometabolic regulation. Here, we present a multivariate framework that applies network analysis and systematic evaluation of pairwise metabolite ratios to a reference dataset of 53 individuals, revealing coordinated patterns of metabolite covariation as well as a hierarchical inter-metabolite architecture. We validate the generalizability and robustness of these findings across four independent MRS datasets, encompassing both longitudinal and cross-sectional samples from 78 humans and 7 rats across five brain regions and three field strengths. Our results show that inter-metabolite relationships are broadly conserved across methodological contexts, while selective axes exhibit graded, network-informed modulation in response to physiological challenges such as caloric restriction or transcranial magnetic stimulation. By moving beyond univariate measures, our multivariate framework accommodates the innate complexity in human brain metabolism and carries translational relevance for biomarker discovery and the development of metabolically targeted therapeutic strategies.

## 1 Introduction

Proton Magnetic Resonance Spectroscopy (1H-MRS) has become an essential tool in neuroscience, offering the capability to non-invasively quantify the concentrations of neurotransmitters and other metabolites in the living human brain [1–3]. This ability has not only advanced our understanding of brain metabolism and function but has also played a pivotal role in translational research, particularly in psychiatry and neurology, where many interventions exert their effects through modulating neurochemical pathways [4–6].

Historically, the broader application of MRS has been limited by technical constraints, especially at clinical field strengths, such as 3 Tesla (T). Challenges have included long acquisition times, the need to pre-select singlevoxel regions of interest, and restrictions on the number of metabolites that can be reliably quantified in a given scan. However, recent technological advancements, most notably the increasing use of ultra-high-field MRS (e.g., 7T), have begun to overcome these limitations [7, 8]. These developments now enable faster acquisition, improved signal-to-noise ratios, and more precise quantification of a broader array of metabolites across brain regions. As ultra-high-field systems become more widely available, the potential of MRS for comprehensive neurochemical profiling is likely to expand substantially [9].

Despite these advances, the vast majority of MRS studies still focus on single metabolite levels, sidelining the interdependencies and coordinated regulation among metabolites. However, many metabolites, such as glutamate and glutamine that share biosynthetic pathways, are subject to co-regulation, and function in concert rather than in isolation. Moreover, the balance between excitatory and inhibitory neurotransmitters like glutamate and gamma-aminobutyric acid (GABA) plays a fundamental role in neural circuit dynamics [10, 11]. Alterations in the glutamatergic-to-GABA ratio have been identified across narcolepsy, autism and several psychiatric disorders [12–14]. In the context of brain tumors, choline-to-NAA as well as choline-to-creatine ratios characterize abnormally proliferative tissue and can be predictive of tumor progression and phenotype [15–18]. Alterations in the NAA-to-creatine ratio are reported across various neurodegenerative and neurological diseases, such as Alzheimer’s disease, multiple sclerosis, or epilepsy, often reflecting neuronal loss or dysfunction [19–21].

Moving beyond quantification of individual metabolites to a framework that jointly considers metabolite relationships might provide a more accurate reflection of neurobiological reality. In addition, less is known about how metabolites, such as lactate, myo-inositol, or N-acetylaspartate, interact with neurotransmitter systems or reflect shifts in cellular metabolism that influence synaptic functioning. An enhanced understanding of these interactions would elucidate the mechanisms by which metabolic processes shape neurochemical balances. From both a biological and translational standpoint, moving toward integrated, multivariate analyses is thus imperative. Although prior work has reported metabolite ratios or dynamic coupling between selected compounds, a systematic mapping of in vivo metabolite interdependencies with network tools has not yet been established.

To this end, we introduce a novel analytic framework that moves beyond conventional univariate MRS analysis and examines how distinct metabolites organize into a metabolite network architecture (see Fig. 1). From a translational perspective, this offers an opportunity to understand brain metabolism as a coordinated system, revealing organisational principles that underlie the relationships between signaling brain metabolites, primarily involved in neurotransmission and synaptic activity, and non-signaling brain metabolites, associated with energy metabolism, structural integrity, and glial function. In this work, we first apply a network-analytical approach to characterize the patterns of co-variation among brain metabolites and uncover their coordinated relationships within the metabolite network. Complementary, we analyze the hierarchical principles across all pairwise metabolite ratios and establish a metabolite ratio architecture blueprint, whose generalizability we evaluate across samples, scanning modalities, brain regions and species. Finally, we evaluate the adaptability of inter-metabolite relationships under biological challenges such as caloric restriction or transcranial magnetic stimulation and contextualize them in relation to the metabolic network structure. Taken together, our approach offers a new systems-level lens on human brain metabolism, with the potential to inform fundamental neurobiological understanding, biomarker discovery and the development of metabolically targeted therapeutic strategies [22–24].

## 2 Results

### 2.1 Distinct Brain Metabolite Connection Profiles

First, to characterize the organizational architecture of brain metabolites and uncover their interdependencies, we applied a network-analytical approach to the levels of signaling metabolites and non-signaling metabolites (see Methods 4). Specifically, we modeled metabolite levels of nine compounds extracted from the posterior cingulate cortex (PCC) using MRS in 53 healthy adults from the 7T reference sample [25] (see 4). The analyzed metabolites comprised both signaling metabolites,namely, glutamate (Glu), glutamine (Gln), gamma-aminobutyric acid (GABA), and aspartate (Asp), as well as non-signaling metabolites including N-acetylaspartate (NAA), myo-inositol (Ins), choline-containing compounds (Cho), glutathione (GSH), and lactate (Lac) (See method section for details). In the network, each node corresponds to a distinct metabolite, while edges represent partial correlations [-1, 1] between pairs of metabolites after accounting for the influence of all other measured metabolites (Methods 4). This approach enabled us to move beyond isolated univariate contrasts and instead quantify the multivariate interdependencies that characterize the metabolic landscape. Robustness of the network was established with a bootstrap approach (see Appendix A.2).

**Fig. 1.**
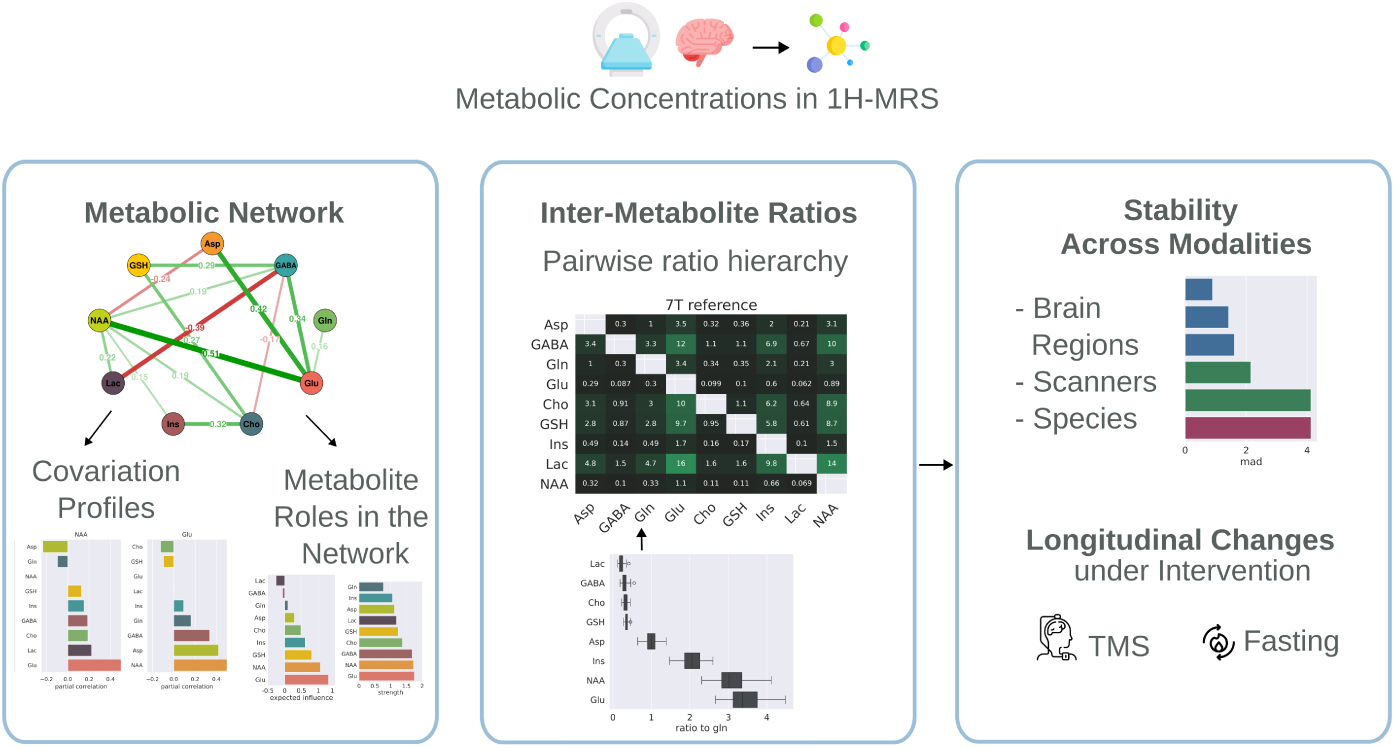
Analysis Overview. We establish a network of metabolites and characterize metabolites by their node strength, expected influence, and individual covariation profile. Complementary, we quantify normative pairwise metabolite ratios and establish a reference inter-metabolite architecture to gain insights into the hierarchical organization of brain metabolism. Finally, we evaluate the stability of this metabolite ratio architecture in independent datasets across different scanners, brain regions and species, and evaluate longitudinal changes under intervention.

To investigate the metabolic network structure, we quantified two complementary nodal metrics. First, we calculated node strength, defined as the absolute sum of edge weights, which captures the overall degree of connectedness for each metabolite. Glutamate, NAA, and GABA emerged as nodes with the highest node strength of up to 1.72 (see Fig. 2b). They are highly integrated in the metabolic network through the magnitude of their associations, both positive and negative, serving as central hub metabolites. On the other end, Gln displayed the lowest node strength (0.77), occupying a rather peripheral position in the metabolic network and indicating that its level is relatively independent of other measured metabolites. Similarly, Ins also showed weak overall connectivity, with all edge weights falling in the range between -0.1 and 0.1 except for the connection to Cho (*r_p_* = 0.3) (see Fig. 2).

Second, we calculated expected influence, which is defined as the signed sum of edge weights, reflecting whether a metabolite’s relationships are predominantly positive or negative. While a positive partial correlation indicates co-occurrence of metabolites, i.e. the presence of one is linked to the presence of the other, a negative partial correlation suggests mutual exclusivity, i.e. the presence of one is linked to the absence of the other. Glu exhibited the largest expected influence (1.3), indicating frequent co-occurrences with other metabolites. Its connection profile is characterized by edge weights of *r_p_* = 0.51, 0.42, and 0.34 to NAA, Asp and GABA, which are at the same time the largest positive connections in the network (see Figure 1c). The second largest expected influence was displayed by NAA (1.06), mainly due to its strong positive connection to glutamate (*r_p_*= 0.51). In contrast to Glu, which exhibits only minor negative partial correlations, NAA also exhibits a strong negative connection to aspartate (*r_p_* = −0.24). Only two metabolites exhibited an overall negative expected influence. In particular, Lac and GABA exhibited a negative expected influence (−0.25 and −0.06), suggesting that their network interactions are more antagonistic and tend to be inversely related to other metabolites. Notably, the partial correlation between Lac and GABA is, at the same time, the strongest negative connection in the network (*r_p_* = −0.39), suggesting a mutual exclusivity in their antagonistic interactions within the metabolic network.

In general, each metabolite displayed distinct connectivity profiles of overlap and segregation with other metabolites (see Fig. 2c and Table 1). The connection profiles of Glu, Ins, and NAA featured the most positive connections, suggesting these metabolites participate in metabolic processes that are broadly co-regulated with a range of others. In contrast, the connectivity profiles of GABA and Gln exhibited predominantly negative associations, consistent with a more reciprocal or antagonistic mode of regulation.

**Table 1.** Edge weights (partial correlations) between the nodes of the metabolic network, as well as node strength and expected influence (EI)

|  | NAA | Ins | Cho | Glu | Gln | GABA | Lac | Asp | GSH | Strength | EI |
| --- | --- | --- | --- | --- | --- | --- | --- | --- | --- | --- | --- |
| NAA | 0.00 | 0.15 | 0.19 | 0.51 | -0.10 | 0.19 | 0.22 | -0.24 | 0.13 | 1.72 | 1.06 |
| Ins | 0.15 | 0.00 | 0.32 | 0.09 | -0.10 | -0.12 | 0.12 | 0.05 | 0.10 | 1.06 | 0.61 |
| Cho | 0.19 | 0.32 | 0.00 | -0.12 | 0.15 | -0.17 | -0.15 | 0.00 | 0.27 | 1.37 | 0.48 |
| Glu | 0.51 | 0.09 | -0.12 | 0.00 | 0.16 | 0.34 | 0.00 | 0.42 | -0.10 | 1.74 | 1.30 |
| Gln | -0.10 | -0.10 | 0.15 | 0.16 | 0.00 | -0.05 | -0.06 | -0.03 | 0.12 | 0.77 | 0.10 |
| GABA | 0.19 | -0.12 | -0.17 | 0.34 | -0.05 | 0.00 | -0.39 | -0.14 | 0.29 | 1.68 | -0.06 |
| Lac | 0.22 | 0.12 | -0.15 | 0.00 | -0.06 | -0.39 | 0.00 | 0.12 | -0.12 | 1.18 | -0.25 |
| Asp | -0.24 | 0.05 | 0.00 | 0.42 | -0.03 | -0.14 | 0.12 | 0.00 | 0.11 | 1.11 | 0.29 |
| GSH | 0.13 | 0.10 | 0.27 | -0.10 | 0.12 | 0.29 | -0.12 | 0.11 | 0.00 | 1.23 | 0.80 |

### 2.2 Inter-metabolite architecture

While network analysis delineates the interdependencies among metabolites in terms of covariation patterns, it does not immediately convey intuitive insights into pairwise metabolite relations, such as their relative abundance. To complement the network perspective, we therefore propose to quantify the relative ratio between metabolite pairs and establish a first normative reference. Evaluating pairwise metabolite ratios not only enables intuitive comparison but also enables us to gain insights into the architectural organization of brain metabolism. Scientifically, shifts in this architec-ture, such as fluctuations in glutamate-to-glutamine or lactate-to-aspartate ratios, can reflect flux through neurotransmitter cycling or energetic pathways, and deviations from normative ratios may signal disruptions in neurochemical homeostasis. Transla-tionally, these metabolite ratios therefore offer the potential to serve as biomarkers in the detection of pathophysiological disturbances in metabolic network organisation.

**Fig. 2.**
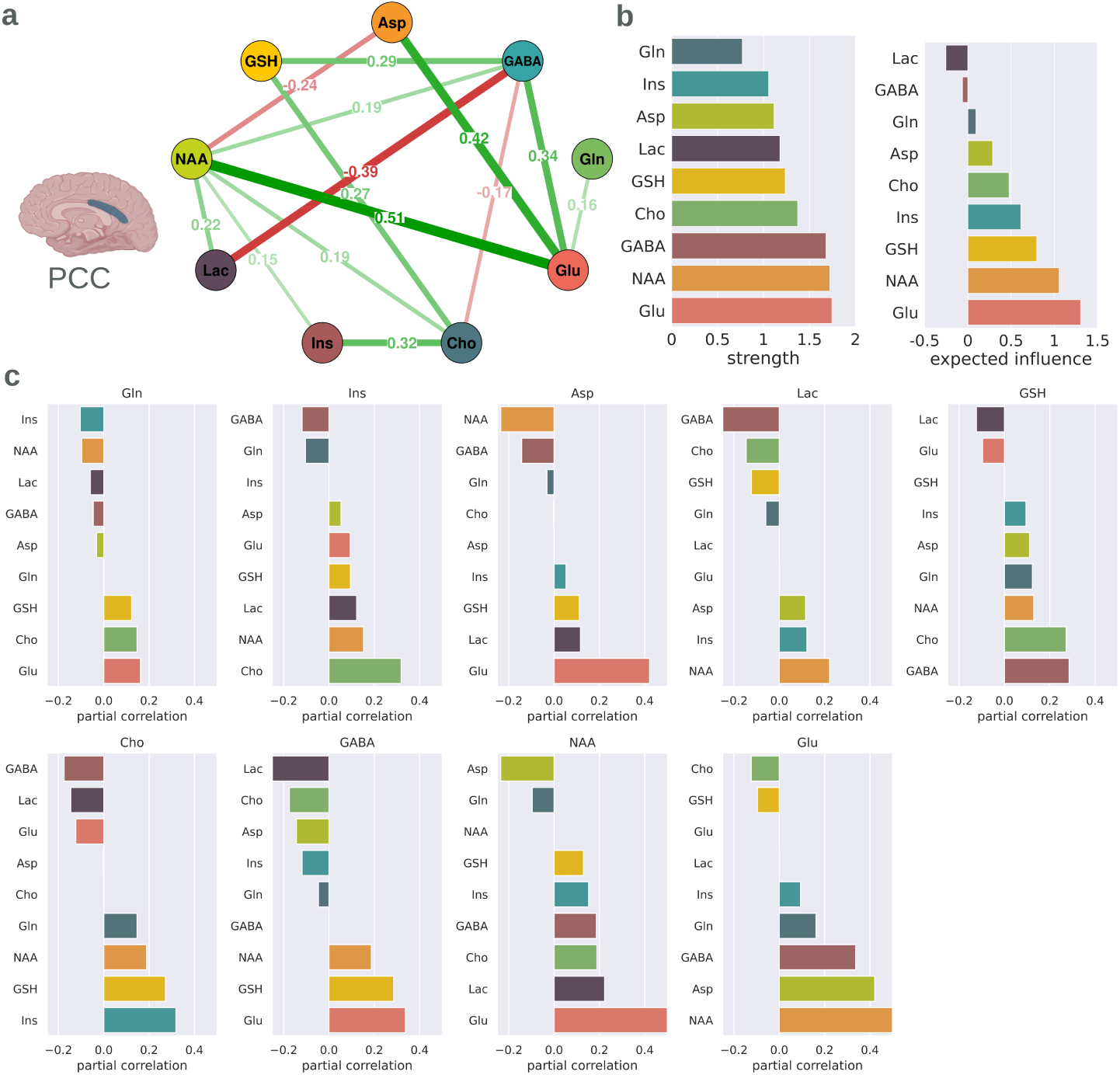
Metabolite Network, Network Metrics and Covariation Patterns for the 7T Ref-erence Sample. **a** The metabolite network, nodes representing one of nine metabolites, while the edges display partial correlations between nodes (partial correlations *<* 0.15 omitted for readability in the network depiction). **b** Metabolite network metrics, ranking all metabolites in terms of strength (overall connection magnitude) and expected influence (sum of connections with respect to the con-nection sign). **c** Connection profiles of all investigated metabolites showcasing their partial correlation within the network.

Visualizing the median metabolite ratios reveals a distinct architectural pattern of inter-metabolite ratios (see Fig. 3). The ratio between Glu and Lac stands out with the largest magnitude, with the level of glutamate being 16 fold of Lac. In contrast, the Glu to NAA ratio is 1.1, indicating that levels of Glu and NAA are almost equal (see Fig. 3b and h). Similarly, while the ratio between Ins and Lac is 10, positioning Ins as manifold in its presence compared to Lac, the ratio from Ins to NAA is 0.66. This systematic quantification and mapping of metabolite ratios provides a normative metabolic landscape for future comparative work. To support the robustness of these findings, we evaluated the stability of the metabolic ratio pattern in independent datasets.

**Fig. 3.**
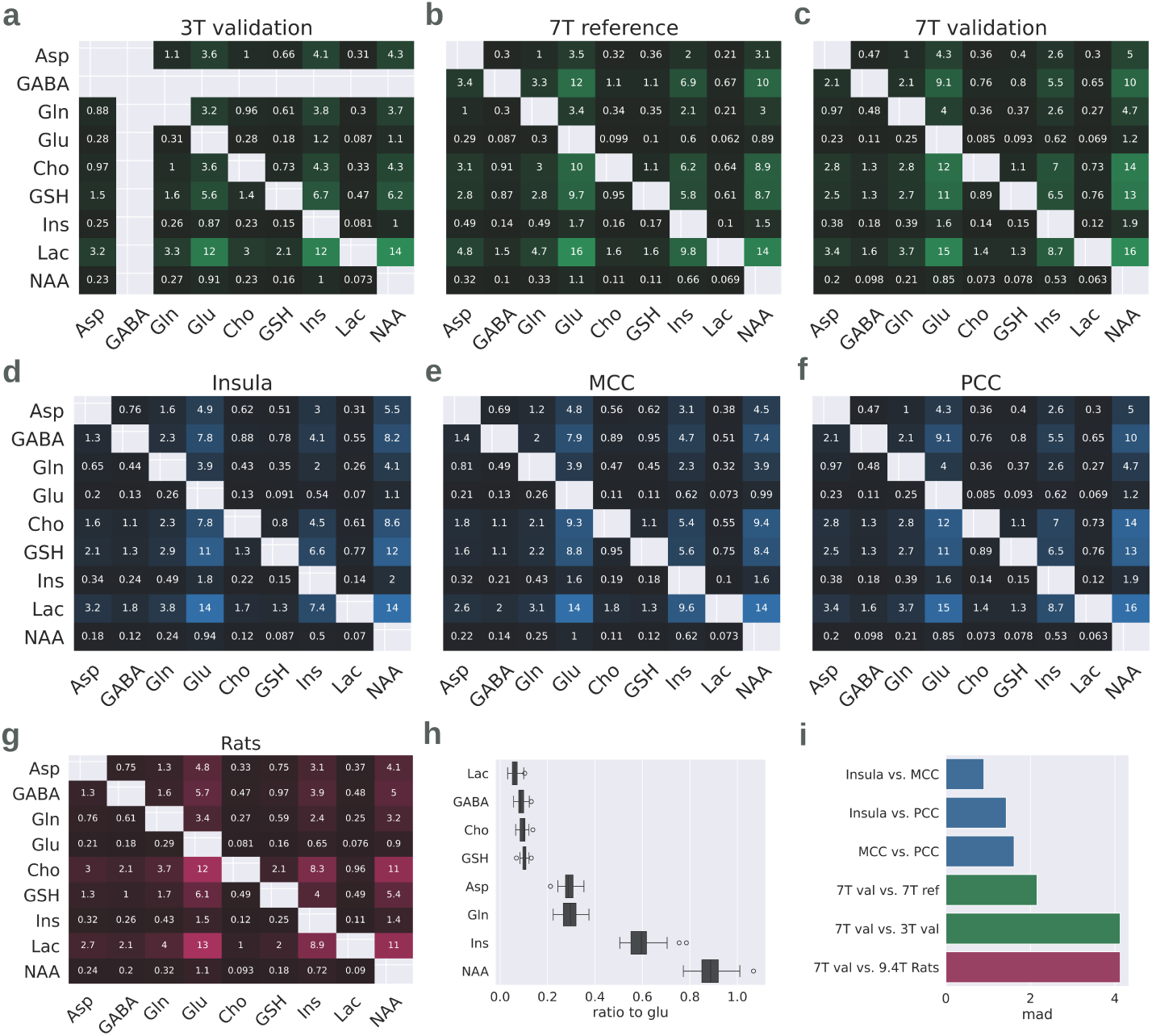
Comparison of metabolite ratio patterns across different modalities. **a-g**. Heatmaps of the pairwise metabolite ratios depict a remarkably similar architecture across different scanners and field strength (green), brain regions (blue) and across species (red) **h.** Relative metabolite ratios, here exemplified from the perspective of glutamate. While NAA level is nearly the same as that of glutamate (0.9), lactate level is only a fraction (0.1). **i.** Quantitative evaluation of the average difference between metabolite ratio patterns of different acquisition modalities, normed against the respective median absolute deviation, i.e. the typical inter-individual fluctuations observed across individuals. We observed negligible differences between different brain regions (around one standard deviation, blue), differences double the usual fluctuations between individuals across the same field strength and brain region (green), and differences of around four median absolute deviations between scanners of different field strengths or in inter-species comparisons.

### 2.3 Stability of the metabolite architecture across modalities, brain regions and species

To assess the generalizability of the metabolic architecture observed in the normative dataset, we validated the observed pattern in different independent datasets. Specifically, we evaluated the pattern across different scanners and acquisition protocols with different field strengths (3, 7, 9.4T), regions-of-interest (PCC, midcingulate cortex (MCC), insula, hippocampus) and across species (humans and rats). Remarkably, despite methodological heterogeneity, the overall pattern and metabolite ratio architecture remained stable (see Fig. 3). For meaningful interpretation, we quantified the difference in metabolite ratios between the different conditions and normalized each difference by the median absolute deviation (mad) observed within individuals for each pairwise metabolite ratio. This approach enabled us to express group differences in a comparable and intuitive unit and provided a measure of divergence from normative patterns (see Fig. 3i).

#### 2.3.1 Stability across scanners and field strengths

We compared the metabolite architecture between datasets acquired on different scanners (see Fig. 3a-c). In particular, we compared the reference dataset (7T reference) to an independent dataset (7T validation), both measuring levels in the PCC at 7T field strength. The average difference in metabolite ratios was 2.15 median absolute deviations, thereby resembling twice the typical level of fluctuation observed across individuals and indicating a high degree of reproducibility even across sites and hardware platforms. In addition, we compared the 7T reference dataset to an independent dataset recorded with 3T field strengths in the hippocampus (3T validation). Here, the differences increased substantially (4.12 median absolute deviations), approaching the magnitude of interspecies variability (see below).

#### 2.3.2 Stability across brain regions

Next, we addressed whether this metabolic architecture was preserved across different anatomical regions using the 7T validation dataset, which offered levels sampled from the insula, MCC, and PCC (see Fig. 3d-f). The regional differences in metabolite ratio patterns were consistently smaller than one standard deviation, i.e. less than typical fluctuations seen across individuals. Notably, the ratio differences between MCC and PCC (1.62 mad), and between insula and PCC (1.42 mad), exceeded the difference observed between insula and MCC (0.90 mad). Our findings indicate that regional metabolite ratios are highly conserved within an individual scanner and acquisition setup.

#### 2.3.3 Stability across species

Finally, we extended our validation to cross-species comparisons, analyzing metabolite ratios in hippocampal samples from a 9.4T rat dataset. Here, the average difference between the human reference (7T PCC) and the rat hippocampus was measured at 4.12 median absolute deviations. Thus, it was comparable in magnitude to the technical differences between field strengths and sites in human data. Although interspecies differences were the largest observed, the hierarchical organization of metabolite ratios remained substantially preserved, indicating translational potential.

Overall, the architectural properties of the metabolic ratios are observably similar. Our quantitative comparison revealed that the difference between different metabolite ratio architectures remained between magnitudes of 1-2 median absolute deviations within different brain regions at the same scanner and different scanners with the same field strengths, i.e. roughly similar or double to common inter-individual fluctuations.

### 2.4 Longitudinal changes in the metabolic ratio architecture following intervention

Finally, we tested whether the organizational principles that structure the metabolic architecture remain stable under biological challenge, or if critical metabolic relationships adapt in response to an intervention.

To this end, we analyzed ultra-high-field (7T) MRS data from the PCC in twelve healthy participants (n=12) at baseline and following a metabolic intervention in the form of a caloric restriction regimen (Buchinger Fasting). To assess the specificity of the observed effects, we further examined three additional datasets, each comprising two longitudinal 3T MRS measurements of five metabolites (NAA, Cho, Glu, Gln, GSH) acquired in the anterior cingulate cortex (ACC): patients with major depressive disorder (MDD) undergoing repetitive transcranial magnetic stimulation (TMS; n=11) or treatment as usual (TAU; n=22), and a healthy control cohort (n=21) assessed without intervention.

Comparisons of inter-metabolite ratios across baseline and follow-up sessions revealed intervention-specific patterns of metabolic reconfiguration. During fasting, the strongest modulation centered on Gln, with the Ins/Gln ratio showing the largest change (*η*^2^ = 0.17), followed by NAA/Gln (*η*^2^ = 0.14) and Glu/Gln (*η*^2^ = 0.13). However, not all Gln-related ratios were affected equally, indicating that specific metabolic relationships are preferentially modulated under physiological challenge.The remaining ratios exhibited small or negligible effect sizes (*η*^2^ between 0.03 and 0.05; Gln/Lac *η*^2^ *<* 0.01). The particularity in the metabolic response was mirrored in the other interventions, albeit with a distinct profile. TMS treatment induced prominent shifts in GSH-related ratios, including Gln/GSH (*η*^2^ = 0.18), Glu/GSH (*η*^2^ = 0.13), and GSH/NAA (*η*^2^ = 0.11). In contrast, effect sizes remained largely minimal or small in the MDD treatment-as-usual arm and in controls, with negligible evidence for global shifts around particular metabolites, underscoring that neither non-specific clinical trajectory nor test-retest variability accounted for the observed network flexibility in the TMS or fasting interventions (see Fig. 4).

Notably, in the fasting interventions, the relationships most susceptible to modulation coincide with the strongest links to the immediate or second-level neighbors in the underlying metabolic network, implicating these edges as critical points of adaptive neurochemical reorganization. The network edges between Ins and Gln and NAA and Gln exhibited the largest negative edge weights, and the edge between Ins and Gln showed the largest positive edge weight. In accordance, the corresponding metabolite ratios showed the largest effect sizes in the Fasting intervention. Moreover, several second-level associations showed large modulation. For example, in the metabolic network, Gln is strongly connected to Ins at the first level and Ins, in turn, to Cho and Asp. Correspondingly, the Ins/Cho and Ins/Asp ratios displayed changes of comparatively high magnitude. An analogous organization was observed in the TMS intervention, albeit constrained by the coverage of only five metabolites. Here, the strongest reconfigurations involved GSH-centered connections, with effect magnitudes largely reflecting their respective network strengths. The main exception was Cho, which, despite being the most strongly connected metabolite in the 7T baseline network, showed only the second-largest modulation following TMS in the 3T MRS data. Overall, these results highlight that adaptive shifts in the ratio architecture do not distribute evenly across all metabolite pairs, but might rather occur on network-privileged axes, providing a mechanistic logic to the brain’s metabolic response to challenge.

## 3 Discussion

This study introduces a network-oriented framework for analyzing the relationship between both signaling and non-signaling metabolites in the human brain. Specifically, we first established a reference metabolic network and analyzed the degree of inter-connectedness for each metabolite, its expected influence within the network as well as its unique covariation pattern. Second, we built on biological evidence showing that shifts in inter-metabolite ratios affect neurophysiological functioning and established an architectural pattern of inter-metabolite ratios. Using a longitudinal dataset of a dietary intervention as an exemplary biological challenge, we then demonstrated the modulability of inter-metabolite ratios within the context of the metabolic network structure.

We found the established metabolic network structure to be aligned with prior neurobiological findings. Glutamate and GABA served as central hubs and displayed strong covariation with multiple other metabolites congruent to their central roles in neural excitatory-inhibitory balance [26–28]. Moreover, in accordance with their inhibitory and excitatory functions, GABA exhibited a predominantly negative covariation pattern, while glutamate exhibited a predominantly positive covariation pattern. Notably, the edge weight between GABA and glutamate was positive and of comparatively large magnitude (0.34), indicating that both metabolites tend to co-occur. This positive linkage likely reflects dynamic homeostatic regulation preserving functional equilibrium. Prior studies suggest that, particularly in aging and disease contexts, reductions in both GABA and glutamate signify overall synaptic and neuronal loss rather than a shift towards net excitation or inhibition [12, 29, 30]. NAA exhibited a similar central position in the network. After glutamate, it showed the highest node strength and expected influence in the network. Its central role is in line with its association with dysfunctional states and disease severity across several neurological disorders [13, 19–21]. Notably, however, while glutamate shows strong positive correlations to several metabolites, NAA’s centrality is mainly defined by its strong positive correlation to glutamate, suggesting strong functional coupling. Lactate exhibited the largest negative expected influence in the network, indicating that its presence is linked to the relative absence of other metabolites. A negative expected influence in the network indicates that higher concentrations of the respective metabolite are systematically associated with lower concentrations of other metabolites, reflecting antagonistic or mutually exclusive metabolic relationships across the network. This is congruent with prior findings where increased levels of lactate may signal a network-wide adaptation to stress, reflecting metabolic derangement or a compensatory response [31]. In contrast to metabolites with high positive or negative expected influence, there were also metabolites with less connectivity within the network. Metabolites with more segregated roles occur rather independently from the presence or absence of other metabolites. Here, glutamine showed the most segregated role, aligning with its specialized function in bioenergetics, i.e. its capacity to respond to fluctuating physiological (bioenergetical) demands [32].

**Fig. 4.**
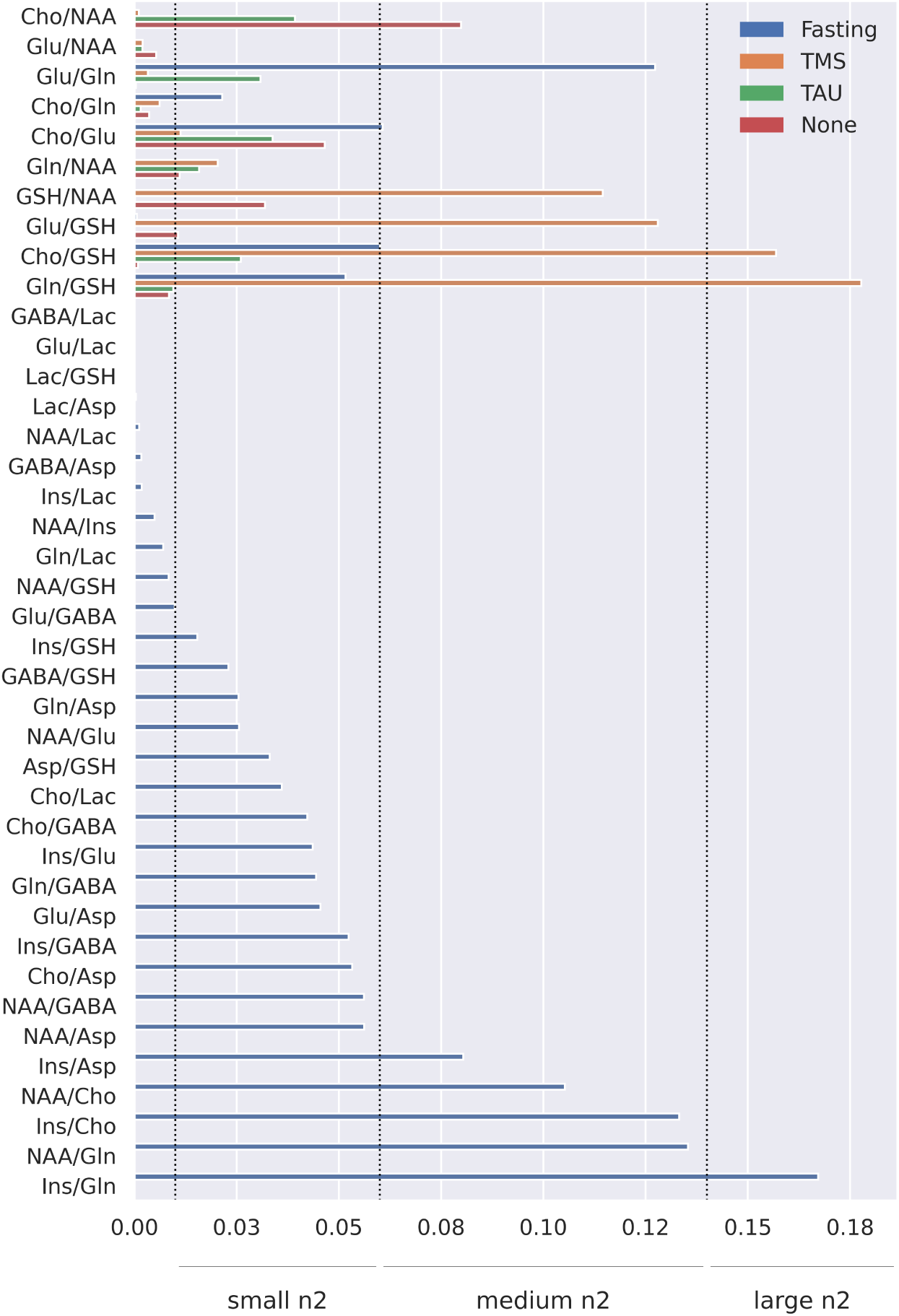
Statistical analyses of longitudinal changes in metabolite ratios. Effect sizes for the comparison of metabolite ratios between baseline measurements and post-intervention: caloric restriction (Fasting; blue) in a cohort of healthy individuals across nine metabolites from 7T MRS. To evaluate sensitivity, we compared these effect sizes with those observed in five metabolites from 3T MRS measurements following transcranial magnetic stimulation (TMS; orange) or treatment as usual (TAU; green) in patients with major depressive disorder, as well as a control group of healthy individuals who received no intervention (None; red).

In sum, the network approach allows for characterization of connectivity and covariation patterns of individual metabolites and may reveal critical control points within the network.While this approach is based on state-of-the-art 7T MRS protocols that quantify multiple metabolite levels simultaneously, we acknowledge that, consistent with common constraints in MRS research, the sample sizes within individual sites were modest (ranging from 12 to 53 participants). This is a relevant limitation for network analyses, which were therefore conducted on our largest available dataset only (n = 53). Future work should build upon this foundation by leveraging larger and more diverse datasets to dissect the dynamic mechanisms and causal directions underpinning metabolite network interactions. Such efforts will not only help identify time-resolved biomarkers and adaptive network responses but also open new horizons for systems-level neurochemical modeling in both health and disease.

Building on prior biological evidence showing that metabolite ratio alterations underlie several pathological states, we extended our analyses from covariation patterns to pairwise metabolite ratios and established a reference metabolite ratio architecture indicating hierarchical patterns across all available metabolites. While different inter-metabolite ratios have been investigated within the context of particular diseases, an encompassing inter-metabolite ratio architecture has, to the best of our knowledge, not been established yet. We used data from healthy controls to establish a first reference metabolite ratio architecture and compared it to multiple MRS datasets from independent sites. To meaningfully interpret the difference between the metabolite ratio architectures of different datasets under constrained sample sizes, we first computed the difference between each inter-metabolite ratio and then normalized it by the respective median absolute deviation found in the reference sample. Thereby, we were able to compare the difference between datasets to the typical fluctuations seen between individuals and convey an intuitive measure of cross-dataset variability relative to normative inter-individual differences seen in male subjects of different age groups. As expected, cohort-specific shifts were observable in some metabolites [33].

However, we found the overall hierarchical pattern to be remarkably consistent across field-strength, acquisition protocols, brain regions and species. This cross-cohort consistency indicates high stability of inter-metabolite relationships and supports their potential utility as potent biomarkers in multicenter and translational research settings. Given the rapidly increasing availability of 7T facilities worldwide, large-scale

investigations of neurometabolic architectures may contribute to the development of generalizable biomarkers for clinical and translational applications. Moreover, future work may explore metabolic diversity in inter-metabolite ratios, which holds promise for precision metabolic phenotyping and may open new avenues for individualized translational interventions.

In a prototypical evaluation of longitudinal MRS data under intervention, we found distinct responsive axes of neurochemical adaptation characterized by unique patterns of shifts in specific metabolite ratios. Rather than interrogating the effects per se, our primary aim was to provide a stringent test of whether key features and connectivity patterns in the metabolite ratio architecture persist, or whether specific relationships are selectively modulated under physiologically meaningful conditions. We found intervention-specific adaptations in two independent datasets and interventions. Both fasting and TMS interventions revealed that only specific relationships within the realm of a responding metabolite are modulated, and that the magnitude of the observed changes was individually nuanced. Moreover, the specific metabolite ratio changes were roughly aligned with the underlying metabolic network structure, indicating that network architecture might be capable of predicting axes of metabolic responsiveness. These findings highlight the multivariate nature of the neurometabolic response to physiological challenge and suggest that network-informed interpretation of metabolite ratios offers a powerful framework for the detection of mechanistic insights into metabolic pathways. Identifying which interactions exhibit the greatest sensitivity to change provides critical insight into the dynamic organization of the braińs metabolic network and delineates potential leverage points through which metabolic or therapeutic interventions may exert targeted influence on neurochemical homeostasis. On the one hand, knowledge of conserved metabolite ratios can guide the preselection of ratio endpoints most likely to respond to a given intervention. On the other hand, future studies may incorporate analyses to outline shifts in ratios beyond the primary targets, thereby capturing broader network-level adaptations that might otherwise remain undetected.

Overall, our findings underscore the promise of metabolite network approaches in revealing insights and finding effective metabolic modulation strategies. As multiple metabolites act in concert to orchestrate brain function and neural states, this multi-variate framework embraces neurobiological reality, thereby offering enhanced analytical potential in the identification of promising candidates for targeted interventions and their biological readouts.

## 4 Methods

To establish a basic metabolic network and evaluate the normative patterns of brain metabolite ratios, we used a reference dataset of 7T 1H-MRS in 53 healthy controls. In this dataset, single voxel spectra were measured in the PCC. Moreover, we validated our findings using four additional datasets acquired across different study sites, scanner hardware, brain regions, and species. Our analyses encompassed nine metabolites, specifically NAA, Ins, Cho, Glu, Gln, Lac, Asp, GSH. For the acquisition protocol details and demographic information of all studies, please see Table 1 and Table 2 in the supplementary materials. Standardized quality control steps were applied at each stage, including spectral preprocessing, peak fitting, and exclusion criteria based on signal-to-noise, linewidth, and fit residuals, all of which were adjusted to the study-specific acquisition protocols to follow current best practices for MRS data reliability (please see Appendix A.1).

### 4.1 Materials

**Table 2.** Datasets overview and demographics.

| Dataset | n | T | ROIs | Sex and Age |
| --- | --- | --- | --- | --- |
| 7T Reference Sample | 53 | 7T | PCC | 34 males; $29.27 \pm 10.83$ years |
| 7T Validation Sample | 12 | 7T | PCC, MCC, Ins | 7 males; $26.17 \pm 4.99$ years |
| 3T Validation Sample | 12 | 3T | Hippocampus | 4 males; $64.2 \pm 10.5$ years |
| 3T Intervention Sample TMS | 11 | 3T | ACC | 6 males; $49.18 \pm 13.61$ years |
| 3T Intervention Sample TAU | 22 | 3T | ACC | 13 males; $41.5 \pm 13.33$ years |
| 3T Intervention Sample Control | 21 | 3T | ACC | 14 males; $39.90 \pm 12.68$ years |
| Rats Validation Sample | 7 | 9.4T | Hippocampus | 7 males; 10–12 weeks |

#### 4.1.1 7T Reference Sample

To establish the metabolic network, as well as normative pairwise normative ratios we leveraged a dataset of 53 healthy subjects[25]. Inclusion criteria were right handedness and no history of neurological and psychiatric illnesses. Exclusion criteria included contraindications to MRI. Data was collected at RWTH Aachen between October 2021 and January 2023. The study protocol was approved by the Ethics Committee of the Medical Faculty of Aachen RWTH. All participants received financial compensation.

#### 4.1.2 7T Validation Sample

For validation, we used 7T 1H-MRS data from a study which investigated the acute effects of caloric restriction on brain metabolism in 12 healthy, normal-weight individuals. Inclusion criteria were normal body weight (BMI 18.5–24.9 *kg/m*^2^), absence of psychiatric or neurological disorders, no history of cardiovascular or metabolic disease, and no current medication affecting glucose metabolism or brain function. Exclusion criteria included smoking, alcohol or substance abuse, contraindications to MRI, pregnancy or breastfeeding, and previously diagnosed diabetes or obesity. Data were collected at Jena University Hospital, Jena, Germany, between October 2024 and November 2024. Individuals were measured prior (baseline) and after a dietary intervention in the form of a caloric restriction regimen (Buchinger Fasting). The follow-up measurement was conducted after 24 hours of fasting. The study protocol was approved by the Ethics Committee of Jena University Hospital and complied with the Declaration of Helsinki. All participants provided written informed consent before participation.

#### 4.1.3 3T Validation Sample

To further evaluate the robustness of our findings, we leveraged a 3 T single-voxel MRS dataset of 12 individuals measuring eight metabolites NAA, Ins, Cho, Glu, Gln, Lac, Asp, GSH. MR spectra were acquired in the left and/or right hippocampus in individuals who had experienced symptoms of transient global amnesia (TGA) at the Charité Universitätsmedizin Berlin, Campus Benjamin Franklin (CBF), Berlin, Germany. Data were collected between January 2020 and May 2025. The study protocol was approved by the Ethics Committee of the Charité, and complied with the Declaration of Helsinki. All participants provided written informed consent before participation.

#### 4.1.4 3T Intervention Sample

To assess sensitivity to longitudinal changes, we leveraged an additional 3 T single-voxel MRS dataset comprising 21 healthy individuals and 33 individuals with major depressive disorder (MDD), of whom 11 received transcranial magnetic stimulation (TMS) and 22 received treatment as usual (TAU). MDD Diagnosis was confirmed by Structured Clinical Interview (SCID). Exclusion criteria included lifetime diagnoses of substance-related disorders, bipolar disorders, and psychotic disorders for depressed patients, any psychiatric disorder for healthy individuals and neurological abnormalities, organic mental disorders, dementia, brain injures or MRI/TMS contraindications for both. MR spectra were acquired in the left and/or right anterior cingulate cortex at the University Hospital Münster, Münster, Germany, pre- and postintervention. Data were collected between October 2020 and May 2023. Measured metabolites included NAA, Cho, Glu, Gln, GSH. The TAU group received standard treatment consisting of combined pharmacotherapy and psychotherapy as determined by their clinicians. The TMS group additionally received 20-30 sessions intermittent theta-burst stimulation (iTBS) over 6 weeks targeting the left dorsolateral prefrontal cortex (DLPFC, Beam F3) at a minimum of 80% of the individual resting motor threshold. The study protocol was approved by the ethics committee of the medical faculty at the University of Münster and complied with the Declaration of Helsinki. All participants provided written informed consent before participation.

#### 4.1.5 Rats

For inter-species comparison, we used data provided by Simicic et al. for in vivo rat brain metabolite levels acquired at 9.4T [34]. Seven adult male Wistar rats (Charles River Laboratories, L’Arbresle, France; age: 10–12 weeks; body weight: 250–320 g) were studied under general anesthesia (1.5–2.5% isoflurane in oxygen). Exclusion criteria included failure to maintain physiological stability during anesthesia, poor spectral quality (linewidth *>* 15 Hz), or motion artifacts during MRS acquisition. Animals were placed on a heated bed to maintain a body temperature of 37.5 *±* 1.0 *^◦^*C. All animal procedures were reviewed and approved by the Committee on Animal Experimentation for the Canton de Vaud, Switzerland.

### 4.2 Statistical Analysis

#### 4.2.1 Network Analysis

Metabolite network structure was characterized using the R package bootnet (add Version). Using the reference dataset, we estimated a non-regularized Gaussian Markov random field network of partial correlation networks, in alignment with best practices [35]. Stability of the network was assessed using a non-parametric bootstrap of 1000 iterations with Benjamini Hochberg correction. To further characterize metabolite roles in the network we evaluated two nodal centrality metrics provided by the package: Node strength, defined as the sum of absolute edge weights, reflecting overall connectivity, as well as expected influence, defined as the sum of signed edge weights, quantifying net positive or negative impact.

#### 4.2.2 Pairwise metabolite ratios

Pairwise metabolite ratios were calculated among all measured metabolites for each individual in the different datasets. To capture population-level patterns, we computed the median for each ratio across participants within a given group, resulting in a median ratio matrix that summarizes the architecture of brain metabolite levels. For comparisons of this architecture between brain regions, scanners, or species, we computed the difference between the respective median ratio matrices across groups. Subsequently, we normalized this difference matrix, by dividing by the standard deviation observed across individuals for each metabolite pair. This approach yields the distance in metabolite ratio architectures in units of typical inter-individual fluctuation and provides an interpretable and intuitive metric. Finally, we report the mean of these normalized ratio differences to provide an overview of how much the metabolic hierarchy diverges between scanners, brain regions, or species.

#### 4.2.3 Statistical Analysis

To systematically compare all pairwise metabolite ratios across the different timepoints in the 7T Validation sample and 3T intervention sample, we conducted an Analysis of Variance (ANOVA) using the Pingouin statistical library in Python, specifying the metabolite ratio as the dependent variable and the timepoint as the predicting factor. Given the modest sample size (n=53), our primary focus was on relative comparisons of effect sizes, quantified as the proportion of explained variance (*η*^2^), rather than on the statistical significance of differences.

## Data Availability

All data produced in the present work are contained in the manuscript.

## 4.4 Acknowledgments

The study was performed in parts by using the research infrastructure of the Werner-Kaiser-Research Center of the Jena University Hospital, Friedrich-Schiller-University Jena.

JR was supported by the LOEWE program of the Hessian Ministry of Science and Arts (Grant Number: LOEWE1/16/519/03/09.001(0009)/98).

SET was funded by the Leistungszentrum Innovative Therapeutics (TheraNova), funded by the Fraunhofer Society and the Hessian Ministry of Science and Art; the Bundesministerium f”ur Forschung, Technologie und Raumfahrt (BMFTR; Federal Ministry of Research, Technology and Space) – 01EO2102 INITIALISE Advanced Clinician Scientist Program; and the REISS foundation.

The financial support by the Austrian Federal Ministry for Digital and Economic Affairs; and the National Foundation for Research, Technology, and Development; and the Christian Doppler Research Association, is gratefully acknowledged.

We would like to acknowledge E.J. Auerbach and M. Marjanska (Center for Magnetic Resonance Research and the Department of Radiology, University of Minnesota, USA) for the development of the STEAM and FAST(EST)MAP sequences for the Siemens platform, which was provided by the University of Minnesota under a C2P agreement.

## 4.5 Author contribution

NO adminstered the project. NO, AG and RL conceptualized the study. IN, RR, NJS, EF, DG, RM, IG, MG, JB, JR, SM, NCG, SET, CU, LVC, AG, NO curated and provided data. RL, NO, MG developed the methods and performed the investigation. GH, WB, PL, RM, AG validated the findings. NO, JR, NCG, JR, SET acquired funding. RL, AG and NO wrote the first manuscript. All authors edited and reviewed the manuscript.

## A Appendix

## A.1 MRS Acquisiton Details

**Table A1.**
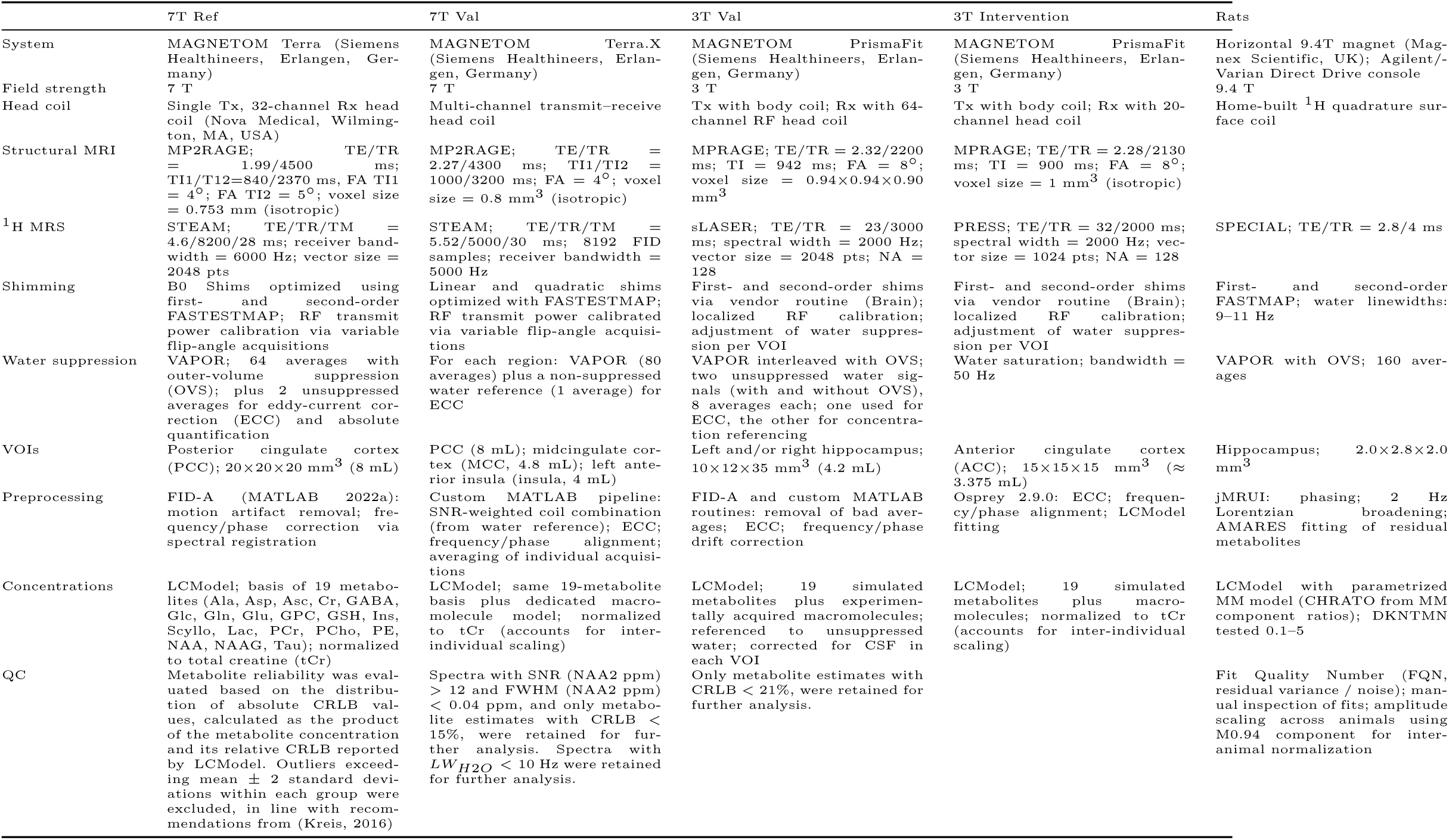
Summary of MRI/MRS acquisition and processing across datasets.

## A.2 Network Stability Analysis

To assess the stability and robustness of the inferred metabolite network structure, we performed nonparametric bootstrapping using the *bootnet* package in R. This involved generating 1,000 bootstrap samples from the original data set of 53 subjects and repeatedly estimating the network for each resample. For every edge in the network, we calculated the mean and standard deviation, as well as the 95% confidence interval (CI) across bootstrap iterations.

Bootstrap analyses indicated a pattern of edge stability consistent with statistical expectations: the precision of edge estimates varied as a function of correlation magnitude and the modest sample size (n=53, see Table A2). Smaller edge weights NAA-Glu, Ins-Cho, Glu-GABA, Glu-Asp, GABA-Lac, GABA-GSH exhibited noticeably wider bootstrap confidence intervals, reflecting greater sampling variability. Importantly, all partial correlations with at least medium effect sizes, i.e. absolute values exceeding 0.3, showed robust stability across bootstrap replications (NAA-Glu, Ins-Cho, Glu-GABA, Glu-Asp, GABA-Lac, GABA-GSH). The confidence intervals for these larger correlations did not include zero, underscoring the reproducibility of the strongest associations in the data. Several additional edge weights, such as those linking NAA-Lac, NAA-GABA, NAA-Asp, and Cho-GSH, demonstrated confidence intervals almost excluding zero, e.g. with upper or lower bounds being *<* 0.06, suggesting that these connections may become more stable and reliably detected with increased statistical power in larger sample sizes. Furthermore, the most pronounced positive (NAA-Glu, Glu-Asp, Glu-GABA) and negative (Lac-GABA, NAA-Asp, Cho-GABA) edge weights not only retained their magnitude but also maintained their relative ordering and directionality with respect to other edges within the network (see Fig. A1). This persistence demonstrates that the key features of the metabolite network consistent even when accounting for sampling variability.

**Fig. A1.**
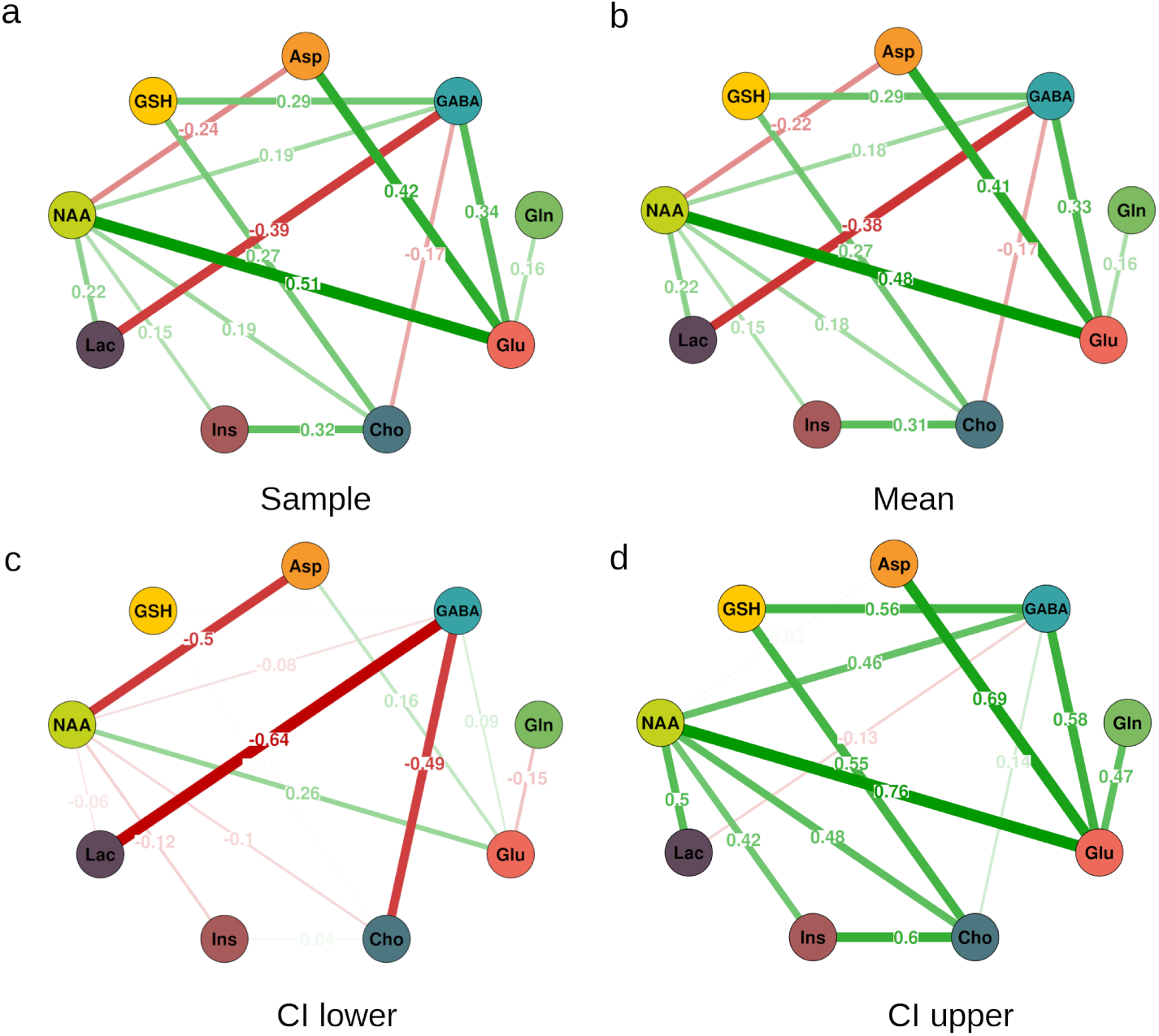
Boostrap results of the metabolite network analysis. Nodes representing one of nine metabolites, while the edges display partial correlations between nodes (partial correlations *<* 0.15 omitted for readability in the network depiction). Stability of the network was evaluated using 1000 bootstraps. **a** Network derived from the sample. **b** Network as defined by the mean of all 1000 bootstraps. **c** Network defined by the lower bound of the 95% confidence interval of the network. **d** Network defined by the upper bound of the 95% confidence interval of the network.

**Table A2.**
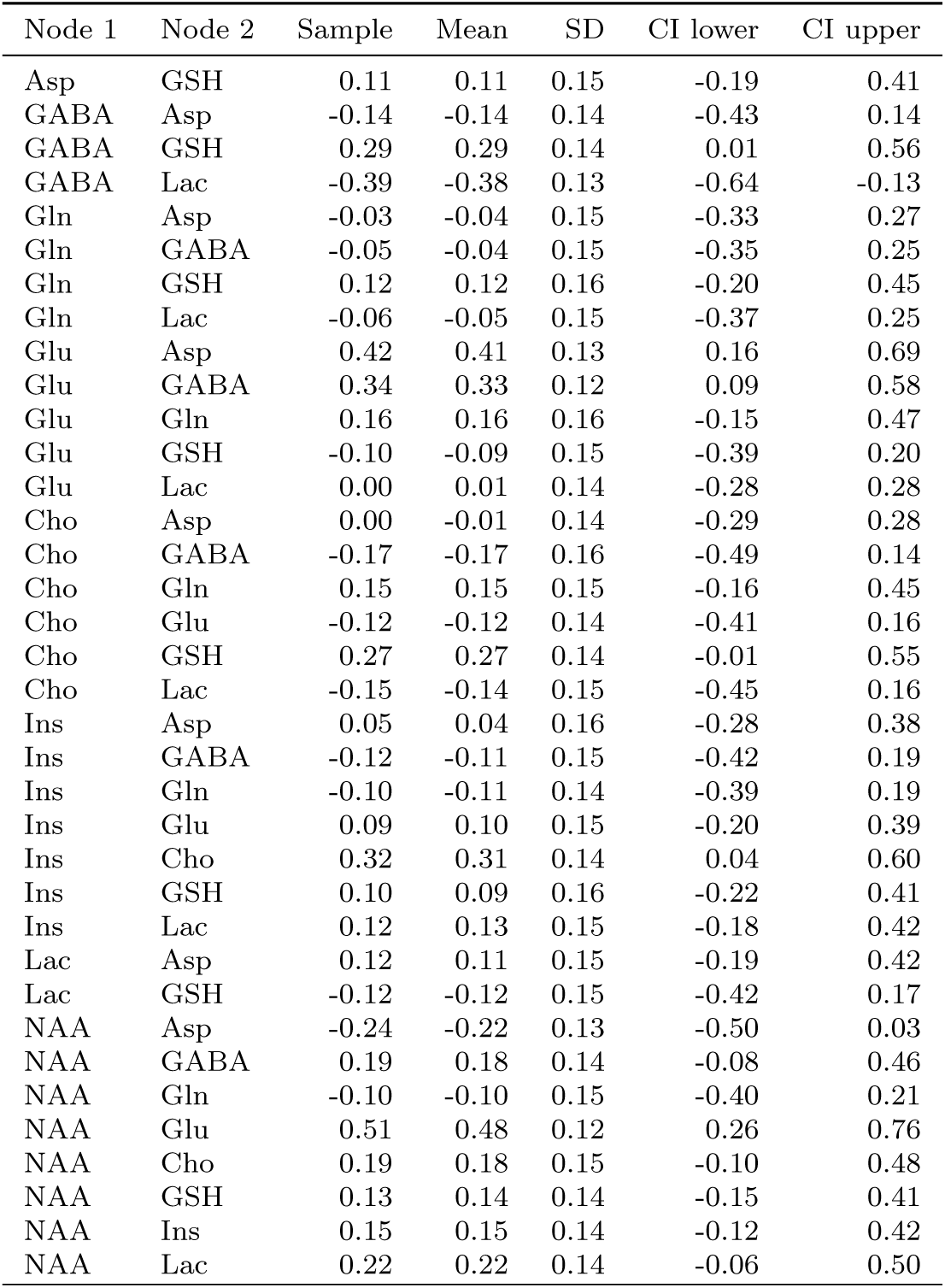
Partial correlations between metabolite pairs (sample, bootstrap mean, SD, and 95% CI).

